# A prospective validation of nomograms based on BC-116 and BC-106 urine peptide biomarker panels for bladder cancer diagnostics and monitoring

**DOI:** 10.1101/2021.12.15.21267846

**Authors:** Lourdes Mengual, Maria Frantzi, Marika Mokou, Mercedes Ingelmo-Torres, Michiel Vlaming, Axel S. Merseburger, Marie C. Roesch, Zoran Culig, Antonio Alcaraz, Antonia Vlahou, Harald Mischak, Antoine G. Van der Heijden

## Abstract

**Purpose:** Non-invasive urine-based biomarkers for bladder cancer (BC) diagnosis and surveillance can potentially improve current diagnostic and monitoring protocols by guiding cystoscopy. Here, we aim to access the diagnostic performance of nomograms based on published biomarker panels for BC detection (BC-116) and monitoring of recurrence (BC-106) in combination with cytology, in two prospectively collected patient cohorts.

**Experimental Design:** 602 recruited patients were screened for presence of BC, out of which 551 were found eligible for further analysis. For the primary setting, urine samples from 73 eligible patients were analyzed from those diagnosed with primary BC (n=27) and benign urological disorders (n=46). For the surveillance setting, 478 eligible patients were considered (83 BC recurrences; 395 negative for recurrence). Urine samples were analyzed with capillary electrophoresis coupled to mass spectrometry and the biomarker score was estimated via a support vector machine-based software.

**Results:** Validation of the BC-116 biomarker panel resulted in 89% sensitivity and 67% specificity (AUC_BC-116_=0.82), similar to the published estimates. The nomogram based on cytology and BC-116 resulted in good (AUC_Nom116_=0.85) but not significantly better performance than the BC-116 alone (P=0.5672). BC-106 biomarker panel showed 89% sensitivity and 32% specificity for surveillance, while improved performance was achieved when a nomogram including BC-106 and cytology was evaluated (AUC_Nom106_=0.82), significantly outperforming both cytology (AUC_cyt_=0.72;P=0.0022) and BC-106 alone (AUC_BC-106_=0.67;P=0.0012).

**Conclusions:** BC-116 biomarker panel is a useful test for detecting primary BC. BC-106 classifier integrated with cytology and showing >95% negative predictive value, might be useful for decreasing the number of cystoscopies during surveillance.

## Introduction

Bladder cancer (BC) has the highest recurrence rate of all cancers. The overall lifetime risk for BC is 2.4%. In Europe, the age-standardized incidence and mortality rates for BC are 11.3 and 3.0 per 100,000 person years, respectively (1,2). In the USA, the respective rate is 10.9 per 100,000 person years, while the annual number of attributable deaths is 2.1 per 100,000 persons (2,3). Most incident BC cases (roughly 75%) are non-muscle invasive (NMIBC) tumors, exhibiting a 5-year recurrence rate of 50-70% and a 10-30% 5-year progression rate to muscle-invasive disease (MIBC), thus necessitating long-term patient monitoring (4). Both the timely detection of primary/ incident BC and efficacious surveillance of BC recurrence are vital for optimal patient outcomes. Cystoscopy remains the gold standard for BC detection (5). However, it is an invasive and expensive approach, having also potential complications while subtle recurrences can be easily missed (6). Furthermore, it is bothersome for patients (7). During the corona pandemic, increased waiting times for cystoscopies, and delayed diagnoses are more prominent. The European Association of Urology (EAU) (8) gives priority to patients with high-risk tumors, whereas recommends postponing cystoscopy by six months for patients with low and intermediate –risk tumors, which might be stressful for a lot of patients. Urine cytology is still the most accurate non-invasive test for BC detection having high sensitivity (84%) in high grade (G3) tumors, but low sensitivity (16%) in low grade tumors (G1), while in experienced hands specificity can reach values over 90% (5). However, in recent large prospective multicenter studies it was demonstrated that the performance of urine cytology in contemporary practice proves to be even lower than previously reported (9). Thus, a clear need for a non-invasive approach for BC detection and monitoring is evident. Liquid biopsy has yielded enormous interest in the field of BC (10) with urine being particularly attractive due to its direct contact with the tumor as well as it can be easily collected in a non-invasive manner (11).

Capillary electrophoresis coupled to Mass Spectrometry (CE-MS) provides high-resolution proteomics profiling data, is characterized by high reproducibility (12) and has been applied for the investigation of peptides and low molecular weight proteins as biomarkers derived from body fluids, especially urine (12,13). Candidate biomarkers can be sequenced and identified by employing CE coupled to tandem MS (MS/MS) and liquid chromatography coupled to MS/MS (LC-MS/MS) proteomics platforms (14), in combination with appropriate bioinformatics tools and algorithms that enable CE-MS data assessment, peptide list extraction and calibration using internal standard peptides (15). Urinary profiling data acquired by CE-MS have been previously explored for detection of BC (16) as well as discrimination of NMIBC from MIBC (17). Moreover, multi-biomarker panels appear advantageous compared to single biomarkers, especially in the context of complex diseases (18). Towards the need for reducing cystoscopies, we previously developed two urinary peptide biomarker panels for: i) non-invasive detection of primary BC (BC-116 biomarker panel) and ii) for monitoring of recurrent BC in patients undergoing surveillance (BC-106 biomarker panel) (19). To develop both biomarker panels, in the above initial study a total of 1,357 urine samples from patients suspected of BC, as well as non-malignant urological controls from five different clinical centers were analyzed through CE-MS. Among the biomarkers were peptides of Small proline-rich protein 3, 14-3-3 sigma protein, CD99 antigen, Apolipoprotein A-I, Fibrinogen α and β, Beta-2-macroglobulin, Basement membrane-specific heparan sulfate proteoglycan core protein and Membrane-associated progesterone receptor component 1. Both biomarker panels exhibited good performance, with the BC-116 biomarker panel reaching 91% sensitivity and 68% specificity, and the BC-106 biomarker panel 87% sensitivity and 51% specificity at the pre-defined threshold. The area under the ROC curve values (AUCs) were 0.87 and 0.75, for detection of primary (BC-116) and recurrent (BC-106) urothelial BC, respectively. When investigating the subpopulation of low/ intermediate-risk patients (NMIBC G1–G2), although in low number of patients (n=26), BC-106 biomarker panel yielded a sensitivity of 89% in detecting recurrent BC (AUC=0.72). Additionally, the negative predictive value (NPV) and positive predictive value (PPV) of BC-116 were estimated at 81.3% and 83.2% (considering a prevalence rate of 63.5%), while for BC-106 at 93.6% and 32.3% respectively (accounting for a prevalence of 21.2%) (19). The predictive value of BC-106 biomarker panel for BC relapse was also evaluated in a follow-up study indicating a prognostic potential of the urinary biomarker panel based on the peptidomics profile in prognosis of BC recurrence [HR: 3.15; 1.73-5.70 (95% CI); P=0.0002] (20).

Based on the above biomarker panels, a pilot assessment showed an added value of the urine biomarker panels when combined with cytology, and thus demonstrated a level of complementarity between the two urological investigations (19). Considering this hypothesis, here we present a multicenter clinical study with the aim to evaluate diagnostic nomograms based on cytology and the BC-116 and BC-106 peptide biomarkers in a prospective setting. To this end, 602 patients were recruited, those with suspicious symptoms or under surveillance that were scheduled for cystoscopic examination to investigate presence of BC.

## Materials and Methods

### Study design and participants

In this study, 602 patients with suspicious symptoms for BC and those patients under surveillance because of a previous BC diagnosis were enrolled at the Radboud University Medical Center in Nijmegen (The Netherlands) and at the Hospital Clinic of Barcelona (Spain). Out of the 602 patients, 18 were excluded due to the presence of other cancers / comorbidities including: i) upper urinary tract cancer (n=14), ii) prostate cancer (n=2), iii) renal cell carcinoma (n=1) and iv) pancreatic carcinoma (n=1). Of the remaining 584 patients, 33 initially defined as positive for BC (based on cystoscopy) were further excluded as histopathological data based on transurethral resection of the bladder tumor (TURB-T) or biopsy were either not available or reported a benign lesion. Therefore, the positive cystoscopy result could not be confirmed. Among the 551 eligible patients, 366 were recruited at the Hospital Clinic of Barcelona and 208 at the Radboud University Medical Center. For the biomarker panel validation, the patients were stratified into two cohorts: a) the primary group including 73 patients positive for primary BC and those with suspicious symptoms scheduled for cystoscopy, and b) the recurrent group, including 478 patients under surveillance, as presented in detail below. In all cases the presence of bladder tumors was confirmed with cystoscopy and histological confirmation (biopsy or TURB-T). Tumor grade and stage were determined according to WHO criteria (21) and Tumor Node Metastasis (TNM) classification (22), respectively. Tumors were classified according to their risk of recurrence and progression into high, intermediate, or low risk based on the EAU guidelines published in 2018 (23). The study was performed in accordance with the Declaration of Helsinki and ethical approval was obtained by local Ethics Committees. Informed consent processes adhered to Institutional Review Board-approved guidelines. Ethical approval for this study was obtained by the Ethics Committee in Medical School of Hannover (ID: 3274-2016). A schematic representation of the study design is presented in **Figure 1**.

**Figure 1.**
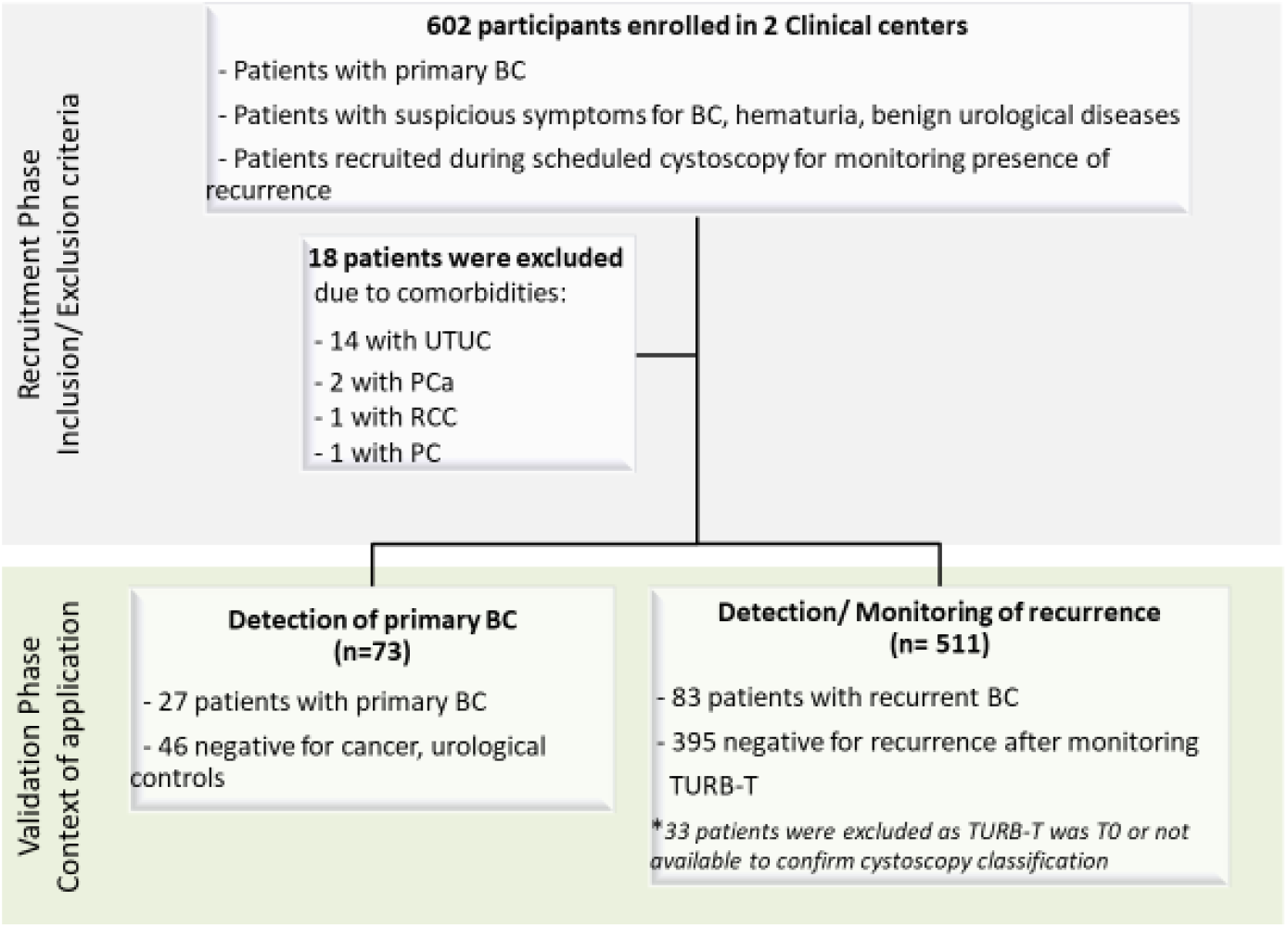
Schematic representation of the study workflow. Patients with primary BC, recurrent BC and urological controls were enrolled at two different clinical centers (Hospital Clinic of Barcelona and Radboud University Medical Center). Patients encountering other comorbidities were excluded from this study (n=18). A total number of 584 participants were considered for further analysis and stratified into two cohorts. The primary cohort was consisted of patients with primary BC (n=73) whereas the recurrent cohort comprised of patients with BC recurrences (n=83) and those negative for recurrence controls (n=395). ***Abbreviations: BC:*** *bladder cancer;* ***UTUC***: *Upper tract urothelial carcinoma;* ***PC***; *pancreatic cancer;* ***PCa***: *prostate cancer;* ***RCC***: *renal cell carcinoma;* ***TURBT***: *trans urethral resection of bladder tumor*.

### Primary cohort

For assessing primary urothelial BC, 73 eligible participants, as per availability, were evaluated. Forty-six patients were enrolled at the Hospital Clinic of Barcelona whereas the remaining 27, were patients undergoing cystoscopy at Radboud University Medical Center. Among the 73 patients, 27 were diagnosed with primary urothelial BC whereas the remaining 46 presented with suspicious symptoms for BC, hematuria, or suffered from other urological diseases (e.g. acute cystitis, nephrolithiasis) and served as urologic controls. At the time of recruitment all patients with primary tumors did not have a prior history of urothelial cell carcinoma and underwent transurethral resection to radically remove the bladder tumor. The cohort characteristics are summarized in **Table 1** and the full list of patient clinical data is given in **Supplementary Table S1**.

**Table 1.**
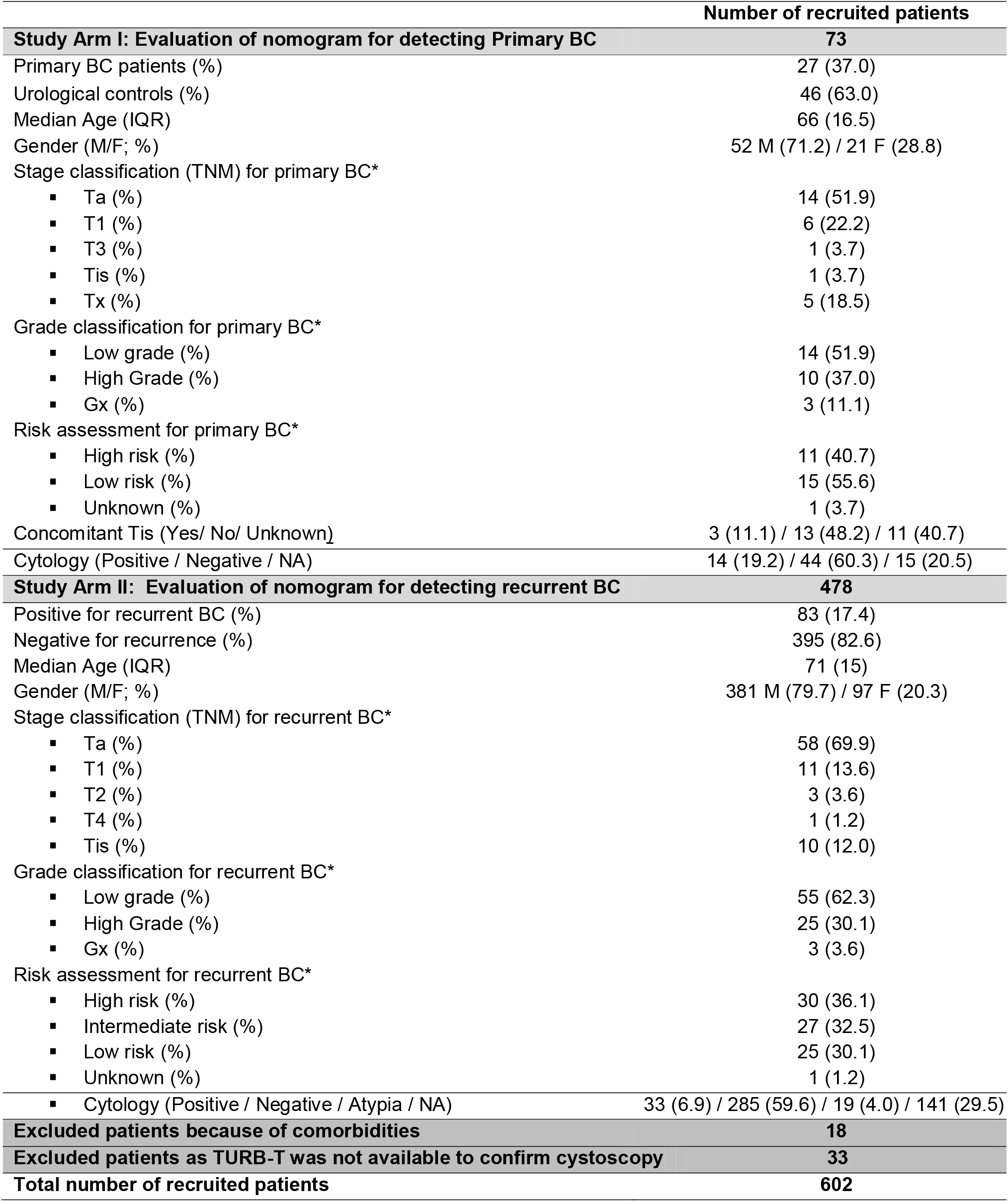
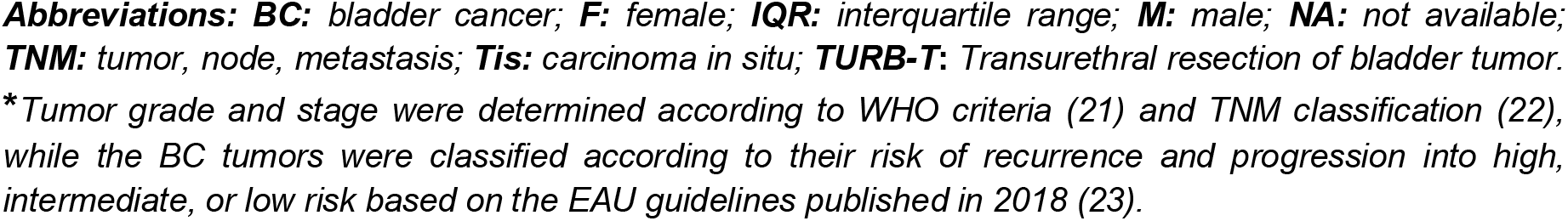
Clinical and demographical characteristics of the patients screened for primary (**Study Arm I**) and recurrence BC (**Study Arm II**).

|  | Number of recruited patients |
| --- | --- |
| **Study Arm I: Evaluation of nomogram for detecting Primary BC** | **73** |
| Primary BC patients (%) | 27 (37.0) |
| Urological controls (%) | 46 (63.0) |
| Median Age (IQR) | 66 (16.5) |
| Gender (M/F; %) | 52 M (71.2) / 21 F (28.8) |
| Stage classification (TNM) for primary BC* |  |
| ▪ Ta (%) | 14 (51.9) |
| ▪ T1 (%) | 6 (22.2) |
| ▪ T3 (%) | 1 (3.7) |
| ▪ Tis (%) | 1 (3.7) |
| ▪ Tx (%) | 5 (18.5) |
| Grade classification for primary BC* |  |
| ▪ Low grade (%) | 14 (51.9) |
| ▪ High Grade (%) | 10 (37.0) |
| ▪ Gx (%) | 3 (11.1) |
| Risk assessment for primary BC* |  |
| ▪ High risk (%) | 11 (40.7) |
| ▪ Low risk (%) | 15 (55.6) |
| ▪ Unknown (%) | 1 (3.7) |
| Concomitant Tis (Yes/ No/ Unknown) | 3 (11.1) / 13 (48.2) / 11 (40.7) |
| Cytology (Positive / Negative / NA) | 14 (19.2) / 44 (60.3) / 15 (20.5) |
| **Study Arm II: Evaluation of nomogram for detecting recurrent BC** | **478** |
| Positive for recurrent BC (%) | 83 (17.4) |
| Negative for recurrence (%) | 395 (82.6) |
| Median Age (IQR) | 71 (15) |
| Gender (M/F; %) | 381 M (79.7) / 97 F (20.3) |
| Stage classification (TNM) for recurrent BC* |  |
| ▪ Ta (%) | 58 (69.9) |
| ▪ T1 (%) | 11 (13.6) |
| ▪ T2 (%) | 3 (3.6) |
| ▪ T4 (%) | 1 (1.2) |
| ▪ Tis (%) | 10 (12.0) |
| Grade classification for recurrent BC* |  |
| ▪ Low grade (%) | 55 (62.3) |
| ▪ High Grade (%) | 25 (30.1) |
| ▪ Gx (%) | 3 (3.6) |
| Risk assessment for recurrent BC* |  |
| ▪ High risk (%) | 30 (36.1) |
| ▪ Intermediate risk (%) | 27 (32.5) |
| ▪ Low risk (%) | 25 (30.1) |
| ▪ Unknown (%) | 1 (1.2) |
| ▪ Cytology (Positive / Negative / Atypia / NA) | 33 (6.9) / 285 (59.6) / 19 (4.0) / 141 (29.5) |
| **Excluded patients because of comorbidities** | **18** |
| **Excluded patients as TURB-T was not available to confirm cystoscopy** | **33** |
| **Total number of recruited patients** | **602** |

**Abbreviations:** **BC:** bladder cancer; **F:** female; **IQR:** interquartile range; **M:** male; **NA:** not available; **TNM:** tumor, node, metastasis; **Tis:** carcinoma in situ; **TURB-T:** Transurethral resection of bladder tumor. \*Tumor grade and stage were determined according to WHO criteria (21) and TNM classification (22), while the BC tumors were classified according to their risk of recurrence and progression into high, intermediate, or low risk based on the EAU guidelines published in 2018 (23).

### Recurrent cohort

For the evaluation of BC recurrence, 511 patients scheduled for follow-up monitoring cystoscopies due to prior history of BC were analyzed in compliance with the EAU guidelines and the European Organization for Research and Treatment of Cancer (EORTC) recurrence and progression scores (23). Among these were 320 patients undergoing cystoscopy at the Hospital Clinic of Barcelona and 181 at the Radboud University Medical Center. In both clinical centers, BC recurrence was confirmed by cystoscopy and histological assessment. Negative score at cystoscopy was used to exclude recurrence and define controls. The recurrent cohort comprised of 83 confirmed BC cases and 395 negative for recurrence controls. Of the 83 relapses, 79 were NMIBC (Ta, CIS, T1) and 4 MIBC (≥T2) cases. The cohort characteristics are summarized in **Table 1** and the full list of patient clinical data is given in **Supplementary Table S2**. Complete information about the clinicopathological tumor characteristics (including risk, stage, and grade) for the previously diagnosed BC tumors is also provided (**Supplementary table S2**).

### Urine collection and cytologic evaluation

All urine samples were collected prior to cystoscopy, if applicable the day before the TURB-T or the day before cystectomy. Voided urine samples were collected in sterile containers and immediately stored at -20°C until further processing. From all patients and controls, only one single sample was included. Without being mandatory, patients were advised to have cytology at cystoscopy or during the period between cystoscopy and surgery. Urine cytology was performed according to Papanicolaou staining and evaluated by expert pathologists in each center blinded to the patient’s clinical history. The results were considered as positive or negative.

### Sample preparation and CE-MS analysis

Urine sample preparation and CE-MS analysis was performed according to previous reports (19,20,24). In brief, 700□µl of each urine sample were diluted with two volumes alkaline buffer containing 2□M urea, 10□mM NH_4_OH and 0.02% SDS (pH 10.5). Thereafter, the samples were concentrated at 1.1 ml using Centrisart ultracentrifugation filters with a cut-off of 20 kDa (Sartorius, Göttingen, Germany) after centrifugation at 3,000g. The filtrate was then desalted using PD-10 columns (GE Healthcare, Munich, Germany) and equilibrated in 0.01% NH_4_OH in HPLC-grade water. The peptide extracts were lyophilized and stored at 4°C until further use. CE-MS analysis was performed using a P/ACE MDQ capillary electrophoresis system (Beckman Coulter, Fullerton, USA) on-line coupled to a MicroTOF MS (Bruker Daltonic, Bremen, Germany), as described previously (19,20). Mass spectral ion peaks representing identical molecules at different charge states were deconvoluted into single masses using MosaiquesVisu software (25). In addition, migration time and ion signal intensity (amplitude) were normalized using 29 internal peptide standards with excretion levels unaffected by any disease state (26). The resulting peak list characterizes each peptide by its molecular mass [kDa], normalized migration time [min], and normalized signal intensity. The detected peptides were annotated, matched and deposited in a Microsoft SQL database (Human Urinary Proteome Database (12,27)) and used as an input in the present study. Accuracy, precision, selectivity, sensitivity, reproducibility, and stability were reported previously (16,24).

### Statistical Analysis

The proportion, mean, standard deviation, median and interquartile range (25th–75th percentiles) estimates were calculated to describe the distribution of the different variables in the patients’ cohorts (summarized in **Table 1**). The biomarker panels’ scores were calculated via the support vector machine (SVM)-based software, namely MosaCluster (version 1.7.0), as previously described (19). Sensitivity and specificity of the biomarker panels were estimated based on the number of correctly classified samples, as defined by cystoscopy. Confidence intervals (95% CI) were based on exact binomial calculations and were calculated along with the receiver operating characteristic (ROC) plots in MedCalc® Version 12.1.0.0 (Mariakerke, Belgium). The area under the ROC curve (AUC) was evaluated to estimate the overall accuracy independent upon a particular threshold (28). The relationship of BC-116 and BC-106 panel with cytology was established using multiple linear regression analyses.

## Results

### Validation of the BC-116 biomarker panel for detection of primary BC

First, assessment of the BC-116 biomarker panel alone was performed to investigate its significance for the BC detection. For the evaluation of the BC-116 biomarker panel, the primary patient cohort was utilized, including 27 patients with primary urothelial BC and 46 patients presenting with non-malignant urological conditions. As shown in **Figure 2a**, the AUC for BC-116 biomarker panel (AUC_BC-116_) was estimated at 0.82 with the 95% CI ranging from 0.71 to 0.90 (P< 0.0001). At the validated cut-off level of -0.27, the sensitivity was estimated at 89% and the specificity at 67%. The classifier correctly classified 24 out of the 27 primary BC cases whereas 15 out of the 46 urological controls were misclassified as BC cases. Considering a prevalence rate of 37%, as estimated on the basis of the participating centers, the NPV was computed at 91% (95% CI 61% to 100%) while the PPV was estimated at 61% (95% CI 33% to 85%). As presented in **Figure 2B**, the BC-116 biomarker panel significantly discriminated urothelial BC cases from controls (P<0.0001, Kruskal-Wallis H test) but also separated controls from cases according to their TNM stage and grade (P< 0.05, Kruskal-Wallis H test; **Figure 2C-D**).

**Figure 2.**
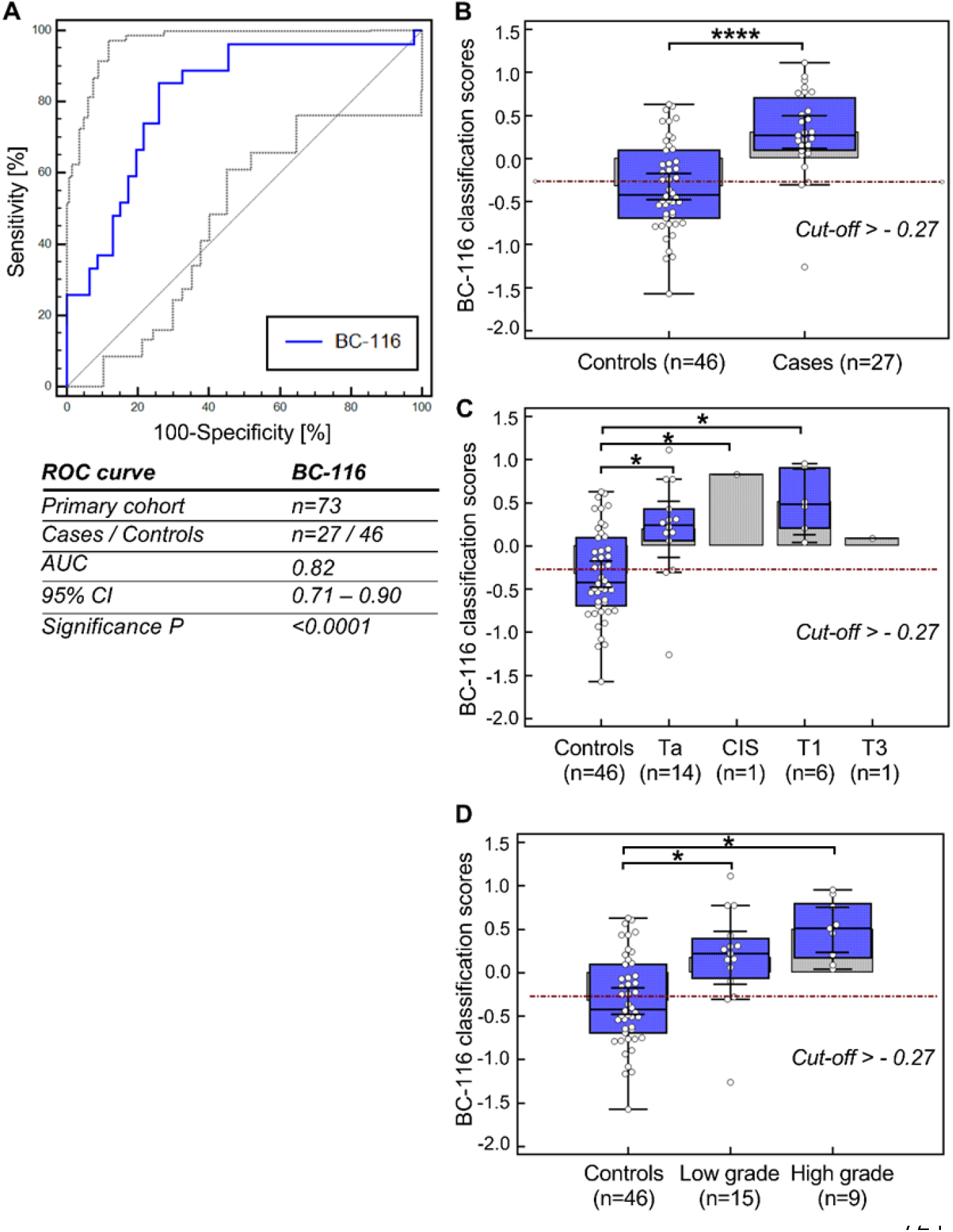
**A**, ROC curve analysis for the urinary biomarker panel BC-116 performed in the primary cohort. The AUC, 95% CI, and P value for the classification of the BC patients are also provided in the table shown in Panel **A. B-D**, Classification scores presented in Box-and-Whisker displaying the level of discrimination between B) the urothelial BC cases and the urologic controls, **C)** BC cases and controls according to their TNM stage, and D) BC cases and controls considering their grade. The average rank differences were significantly different (****P≤ 0.0001 and *P<0.05) as defined with Kruskal-Wallis H test.

### Comparative assessment of BC-116 with cytology

For 58 (out of 73) patients of the primary cohort, data from the cytologic evaluation of the urine samples were available (**Supplementary Table 1**). Sensitivity of cytology for detecting primary BC was estimated at 42.3% while sensitivity of the classifier was 88.5% in this subset of patients. Twelve of the 26 primary BC cases were detected by the BC-116 biomarker panel but were missed by urine cytology. Notably, out of these 12 patients, eight were bearing low grade BC tumors (TaG1) but were also three patients with high grade NMBC (T1G3) and one with MIBC that were missed by cytology and detected by the BC-116. Three cases that were not detected by the BC-116 panel were also missed by urine cytology. These were all patients bearing low grade BC (TaG1). Along these lines, the NPV was higher for the BC-116 classifier (91.2%, 95% CI 60.9 – 99.7) than that of the urine cytology (73.2%, 95% CI 49 – 90.2). Nevertheless, PPV was higher for urine cytology (77.9%, 95% CI 26.7 – 99.2) than for the BC-116 classifier (61.3%, 95% CI 31.4 – 86.0). Specificity of cytology was higher than that of the BC-116 panel (93.7% and 67.2% respectively). Multivariate analysis showed that BC-116 biomarker panel performed significantly better than cytology in primary BC detection, presenting an AUC value 0.84 (95% CI 0.72-0.92) for the classifier and 0.68 (95% CI 0.55-0.80) for cytology (P=0.0195 for pairwise comparison) (**Figure 3A**).

**Figure 3.**
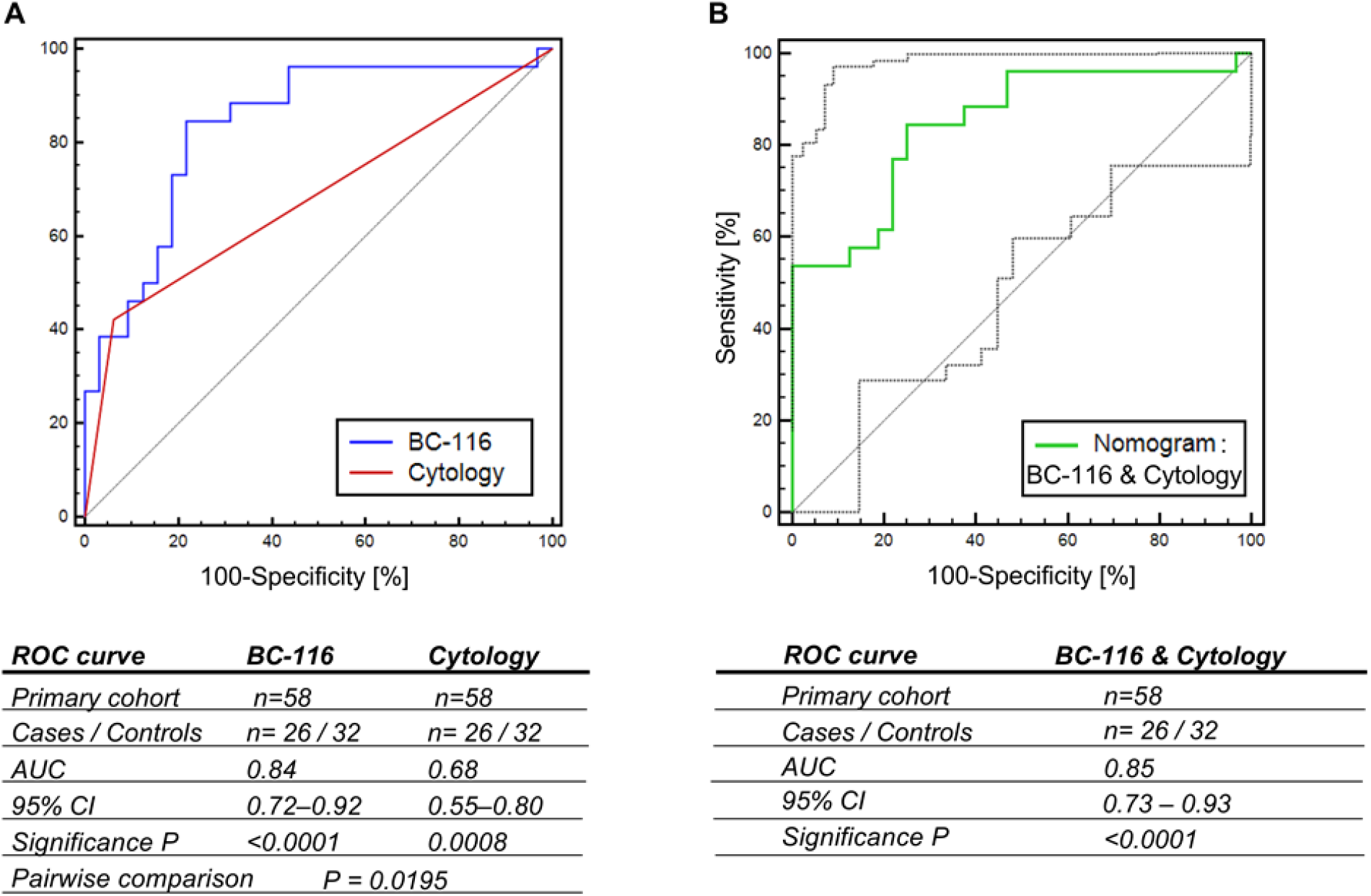
**A**, ROC curves for the urinary BC-116 biomarker panel and the cytology as performed to those patients from the primary cohort with available cytological data. **B**, ROC curve for the integrative nomogram including the BC-116 biomarker panel and the cytology. The AUCs, 95% Cis, and P values are provided in the respective tables.

### Integrative nomogram including BC-116 and cytology for detecting primary BC

An integrated nomogram based on the BC-116 based urinary biomarker panel and cytology was also examined in the primary patient cohort. As shown in **Figure 3B**, the AUC for the integrative nomogram was estimated 0.85 (0.73-0.93; 95% CI; P< 0.0001) only slightly but not significantly higher than the BC-116 alone (0.84; P= 0.5672) yet significantly higher than cytology alone (0.68; P= 0.0016). At the optimal cut-off level of 0.39, the sensitivity was estimated at 84.6% and the specificity at 75%. The integrative nomogram correctly classified 23 out of the 26 primary BC cases whereas 9 out of the 32 controls were misclassified. Considering a prevalence rate of 37%, as estimated on the basis of the participating centers, the NPV was computed at 89.5% (95% CI 60% to 99%) while PPV was estimated at 66.6% (95% CI 35% to 90%).

### Validation of the BC-106 biomarker panel for detection of recurrent BC

The performance of the BC-106 biomarker panel for detection of bladder cancer recurrence was evaluated in 478 patients. Among those, were 83 patients bearing a histopathologically confirmed BC recurrence and 395 controls negative for recurrence. As presented in **Figure 4A**, the AUC was estimated at 0.67 with the 95% CI ranging from 0.63 to 0.72. At the previously established cut-off of -0.63, sensitivity was estimated at 90.4% whereas specificity at 29.1%. This means that 75 out of the 83 recurrences were correctly classified as positive by the BC-106 biomarker panel. However, the number of correctly classified negative controls was only 112. Accounting for a prevalence of 17.4% in the investigated population, the NPV was estimated at 93.2% (95% CI 73.3% - 99.6%) while the PPV was calculated at 21.1% (95% CI 11.7% to 33.4%).

**Figure 4.**
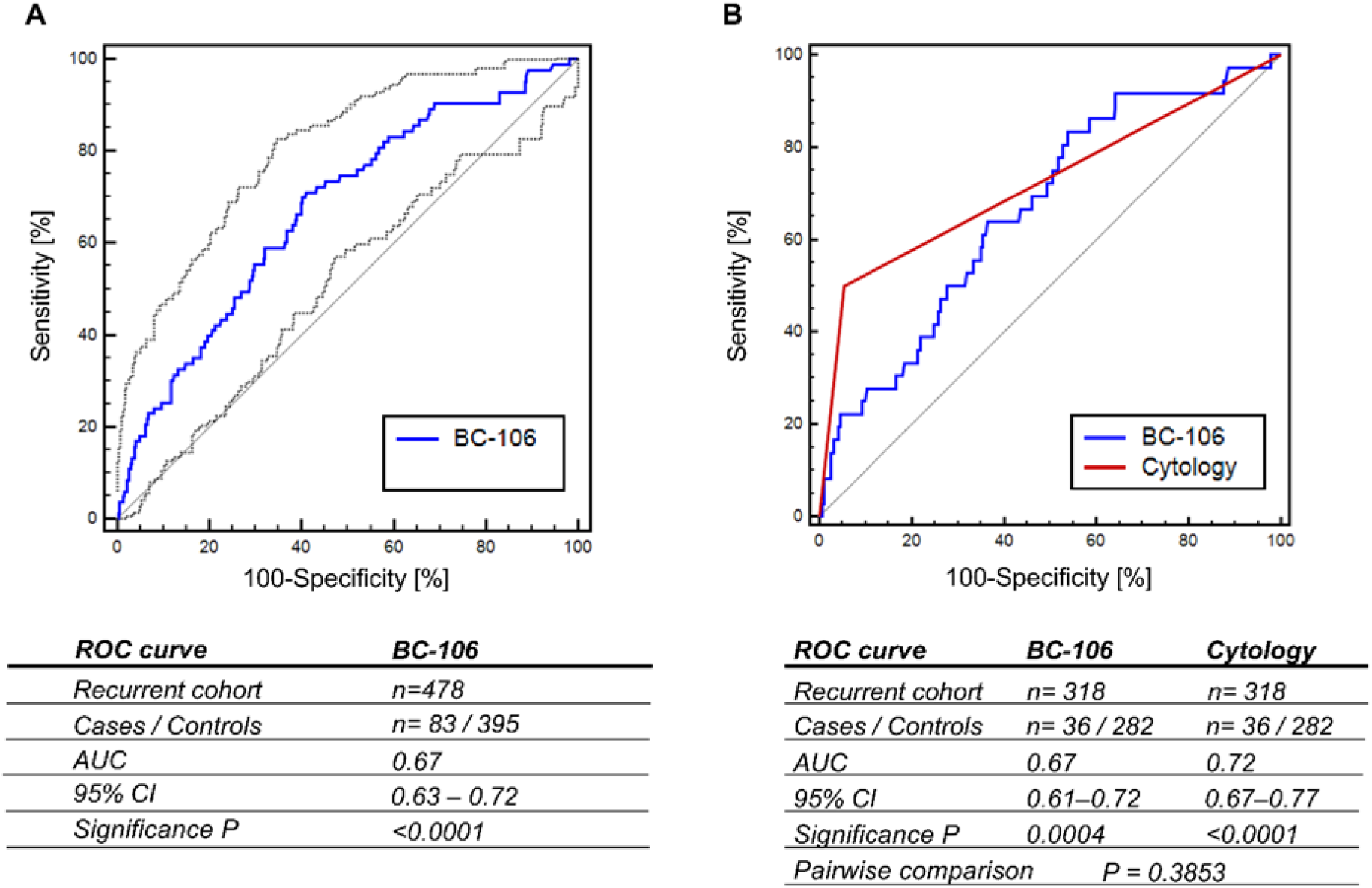
**A**, ROC curve for the BC-106 urinary biomarker panel, consisting of 106 peptides as performed in the recurrent cohort. **B**, ROC curve analysis for the urinary BC-106 biomarker panel as performed in those patients from the recurrence cohort with available cytological data. The AUCs, 95% Cis and P values are provided for the classification of recurrent BC patients.

### Comparison of BC-106 biomarker panel performance with urine cytology

Urine cytology was performed in 318 patients of which, based on cystoscopy, 36 were positive for recurrence cases and 282 were negative for recurrence controls. Sensitivity of the BC-106 biomarker panel for detecting recurrent BC was higher than the one of cytology (91.6% and 50.0% respectively) in this subset of samples. However, the specificity BC-106 biomarker panel was lower than the one of cytology (31.2% for the classifier and 94.7% for cytology). Based on the ROC curve analysis, the estimated AUCs were reported to be comparable between the two tests [AUC for BC-106 was 0.67 (0.61 to 0.72; 95% CI), while AUC for cytology was 0.72 (0.67 to 0.77; P for difference between the 2 tests =0.3853), (**Figure 4B**).

#### Integrative nomogram including BC-106 and cytology for detecting recurrent BC

An integrative nomogram based on the BC-106 biomarker panel and cytology was further validated in 318 patients from the recurrence cohort, for which the results from the cytological examination were available. An improved diagnostic performance was observed when the BC-106 classifier was integrated with urine cytology (AUC = 0.82, 95% CI 0.77-0.86, P< 0.0001), **Figure 5A**. At the optimal cut-off of 0.06, sensitivity was estimated at 86% whereas specificity at 61%. Out of the 36 recurrent BC cases, 31 were correctly classified as positive by the integrative nomogram and out of 282, 172 patients were correctly classified as controls (negative for recurrence). Accounting for a prevalence of 17.4% in the investigated population, the NPV was estimated at 95.4% (95% CI 74.9% - 99.9%) while the PPV was calculated at 31.7% (95% CI 11.3% to 59.1%). The nomogram was found of significantly improved performance in comparison to cytology alone [AUC_NOM106_ was 0.74 (0.69 to 0.78; 95% CI) while AUC for cytology was 0.72 (0.67 to 0.77; P=0,0022)], but also compared to the BC-106 biomarker panel alone [AUC for BC-106 was 0.66 (0.61 to 0.72; 95% CI; P= 0.0012)]. As presented in **Figure 5B**, the integrative nomogram significantly discriminated recurrent BC cases from negative for recurrence controls (P= 0.0005, Kruskal-Wallis H test), but also separated patients with recurrent BC from those negative for recurrence (controls) according to their TNM stage and grade (P< 0.05, Kruskal-Wallis H test; **Figure 5C-D**).

**Figure 5.**
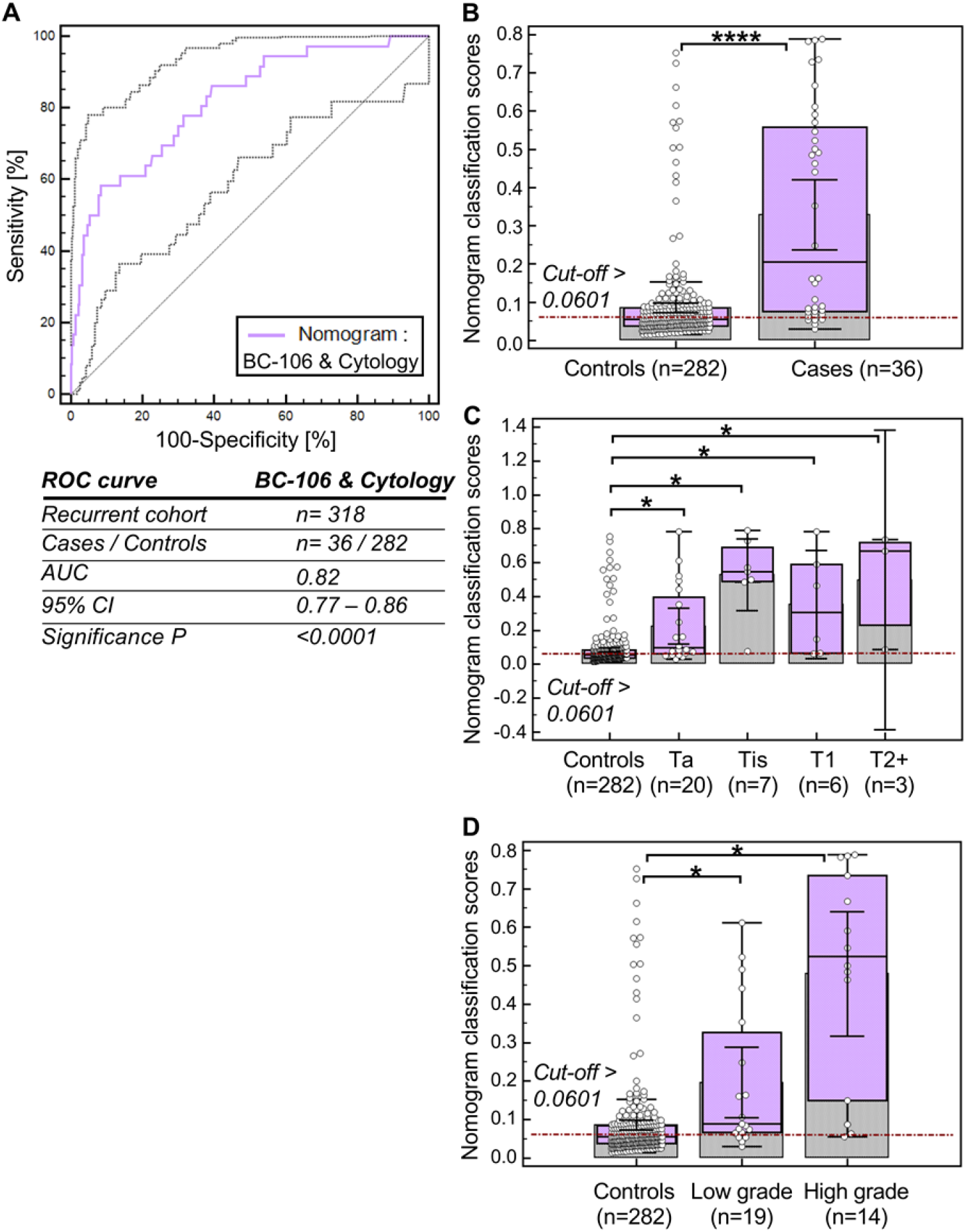
**A**, ROC curve analysis for the integrative nomogram including the urinary BC-106 biomarker panel and cytology, as performed in 318 patients (consisted of 36 positive and 282 negative for recurrence controls) with available cytological data from the recurrent cohort. The AUC, 95% CI, and P value for the integrative nomogram are provided in the table in panel A. **B-D**, Classification scores presented in Box-and-Whisker displaying the level of discrimination between **B)** the recurrence BC cases and the negative for recurrence controls, **C)** the recurrence BC cases and the negative for recurrence controls among stage, and **D)** the recurrence BC cases and the negative for recurrence controls among grade groups. The average rank differences were significantly different ((****P≤ 0.0001 and *P= 0.05) as defined by Kruskal-Wallis H test.

### Reducing the number of follow-up cystoscopies by using the Nomogram BC-106/ cytology

In the participating clinical centers of the study, recurrence was detected in 15.6% of all follow-up cystoscopies while 84% of the patients previously diagnosed with BC were without recurrence at the time of follow-up cystoscopy. At the cutoff point value of 0.06 of the integrative nomogram sensitivity was 86% whereas specificity at 61%. With a very high NPV value (estimated at 95.4%) based on the nomogram, 172 out of 282 patients truly did not bear any recurrence, while only 4 out of the 36 cases would have been missed. All four patients were carrying a low grade BC (TaG1). Based on this, and assuming that patients who present a negative classification based on the integrative nomogram (n=172) will not undergo a cystoscopy, more 60% of all cystoscopies could be prevented, at the cost of 14% of low grade recurrences remaining undiagnosed.

## Discussion

Within the last decade several urinary biomarkers have been developed for BC detection and monitoring (summarized in (29,30)), including among others various promising proteomics and peptidomics biomarkers identified by mass spectrometry (31). To date, none of these biomarkers have replaced cystoscopy as the gold standard for diagnosis and surveillance or has been accepted for diagnosis or follow-up in routine practice or clinical guidelines (5,28). The lack of validation studies presents a challenge in the expanding field of urinary biomarkers, thus delaying their uptake in the clinical setting. In the present prospective study two unique urinary peptide-based biomarker panels that were previously developed for detecting primary (BC-116 panel) and recurrent urothelial BC (BC-106) based on CE-MS proteomics analyses (19) were prospectively validated.

The BC-116 biomarker panel initially developed for detection of primary BC (19) demonstrated excellent reproducibility in this prospective cohort study. BC-116 exhibited good performance (AUC_BC-116_ 0.82) that was comparable to the one previously described (AUC 0.87) (19). Furthermore, BC-116 sensitivity and specificity (89% and 67% respectively) were similar to those previously reported (91% sensitivity and 68% specificity) (19). In this prospective study, BC-116 again performed better than cytology and successfully classified the majority of the BC cases (panel: 24/27 patients; cytology: 11/27 patients). Cytology missed 58% of the tumors, similarly to other studies (32) and did not identify any BC cases that were not previously identified by the BC-116 classifier. Interestingly, 82% of the cases that were missed by cytology were correctly classified by the panel, while three cases that were missed by BC-116 were all low grade BC. The negative predictive value of the classifier was also higher than cytology’s (91% versus 73%). Additionally, an integrative nomogram including BC-116 and cytology was investigated, showing, however only slightly increased performance over the BC-116 alone. These results provide a strong argument for using BC-116 instead of cytology. Considering that CE-MS is validated platform that is applied for routine diagnostics, BC-116 biomarker panel could be potentially applied in clinical practice even as an adjunct to cystoscopy to detect primary or missed tumors. Yet, a cost-effectiveness analysis is required to demonstrate economic efficiency.

For detection of recurrence, the BC-106 panel exhibited similar performance than urine cytology, lower than previously reported (19). The observed reduced performance is mostly attributed to the misclassification of almost half of the negative for recurrence patients. The reduced specificity of BC-106 for detecting recurrent BC could be attributed to differences in the study population and the demographics of the patients. In the initial study, patients with a minimum of one year recurrence-free follow up were investigated, while in this prospective setting, any patient during the regular BC monitoring setting was included, mainly NMIBC (>98%). Importantly, BC-106 showed complementary value with cytology. This fact was already previously observed and has now been replicated in this study. Based on this, the previously reported integrative nomogram including BC-106 and cytology, was also assessed, demonstrating significantly increased performance compared to both cytology alone and the BC-106 alone.

Based on the performance estimates, accounting for NPV values of >90% for primary and >95% for surveillance, the availability of highly sensitive urine peptide markers, in combination with cytology allows for the reduction of the number of follow-up cystoscopies as in the clinical practice only patients with a positive test will undergo cystoscopy, whereas for those with a negative test cystoscopy could be postponed/ skipped (33). The integrative nomogram, including BC-106 classifier and cytology, ruled out 172 of the 282 samples, theoretically reducing the number of follow up cystoscopies by factor 2. Based on the integrative BC-106 model, approximately 14% of the BC patients at follow-up were incorrectly misclassified as not having recurrence, but these were all low grade recurrent BC. Yet, even cystoscopy which is the gold standard misses up to 15% of the papillary and up to 30% of the flat lesions (34). These findings along with the high NPV of the integrative nomogram further supports that a voided urine sample can rule out BC with high confidence; thus reducing the number of cystoscopies and minimizing patients’ discomfort.

Both BC-116 and BC-106 have similar performance as other FDA-approved and/or currently under investigation urinary tests. To date, there are six urinary tests approved by the FDA for clinical use in conjunction with cystoscopy (29,30). Among these, NMP22 Bladder Cancer ELISA test, NMP22 BladderChek point-of-care test and UroVysion have FDA approval for diagnosis and surveillance whereas immunocyte (UCyt+), BTA-TRAK, and BTA-STAT have been approved for BC surveillance following the diagnosis of a primary tumor (29). Several meta-analyses studies have been conducted to evaluate the performance of these panels and a remarkably broad variation in the range of the sensitivity, specificity, NPV and PPV has been observed (30). For instance, in a meta-analysis study of NMP22 BladderChek it was shown that for detecting BC, the pooled sensitivity of the test was 56%, specificity was 88% and the AUC value was 0.83 (35). Similarly, in another study evaluating the utility of UroVysion, it was shown that the overall sensitivity, specificity, PPV, and NPV in detecting BC were 61.9%, 89.7%, 53.9%, and 92.4%, respectively (36). The performance rates of BC-116 and BC-106 panels are at least as good as the FDA-approved tests. However, considering potential confounding factors and the significant variations in the clinical setting, a direct comparison of our biomarkers with the FDA-approved based on the data currently available is complicated and difficult to be accomplished.

A high NPV is important when the panel has a negative score and represents a point of reference to spare unnecessary cystoscopies. This value is critical especially in high-grade disease where a negative biomarker test could result in a missed cancer, thus having detrimental effects with respect to disease progression (30). In the present study, the layout of the clinical settings closely represents a “real-clinical setting” situation. Considering the prevalence rate of each cohort in this study, the NPV was computed at 89.5% for the integrative nomogram of BC-116 classifier and cytology for detecting primary BC and at 95.4% for the integrative nomogram of BC-106 and cytology for detecting BC recurrence, comparable to the previous study (19). Thus, both panels demonstrated high NPV values that may offer benefit to patients with high-risk disease (either with hematuria at primary diagnosis or under surveillance).

One of the limitations of this study is that potential confounding factors such as clinical treatments before the last recurrence were not available for all patients and thus were not considered. Nevertheless, as shown in the initial study, prior treatment did not affect the urinary peptidomic profiles (19). As another limitation of our study, follow-up data were not accessible for all patients. Thus, correlation analysis of false positives cases with later recurrences was not feasible. Therefore, no conclusion can be made on false-positives of BC-116 and BC-106 biomarker panels since they could be attributed to early detection of subclinical recurrence that could not be detected by cystoscopy. This has been previously described for other FDA-approved urinary biomarkers (37). Furthermore, no investigations were performed in patients that had a positive test but a negative cystoscopy to rule out the presence of upper tract tumors.

Collectively, this prospective study provides evidence on the clinical relevance of the BC-116 and BC-106 biomarker classifiers for BC screening and monitoring. As supported by the presented results, such non-invasive biomarker panels can facilitate BC diagnosis (BC-116) and can be applied as adjuncts to cystoscopy in combination with cytology (BC-106) to reduce the number of follow-up cystoscopies as well as patient discomfort and financial burden.

## Data Availability

All data produced in the present study are available from the corresponding author upon reasonable request.

## Acknowledgments

This work was supported in part by BioMedBC (752755; H2020-MSCA-IF-2016), funded by the European Commission.

## Supplementary Files

**Supplementary Table S1**. Clinical and demographical data for the 73 samples from patients with primary BC.

**Supplementary Table S2**. Clinical and demographical data for the urine samples (n= 478) from recurrent BC patients.

**Supplementary Table S3**. List of scoring data for 73 patients with primary BC and 478 patients that were screened for recurrent BC.

